# Corticosteroid Use in Severely Hypoxemic COVID-19 Patients: An Observational Cohort Analysis of Dosing Patterns and Outcomes in the Early Phase of the Pandemic

**DOI:** 10.1101/2020.07.29.20164277

**Authors:** Omar Rahman, Russell A. Trigonis, Mitchell K. Craft, Rachel M. Kruer, Emily M. Miller, Colin L. Terry, Sarah A. Persaud, Rajat Kapoor

**Author notes:** Corresponding Author: Omar Rahman, Department of Pulmonary and Critical Care Medicine, Indiana University School of Medicine, 1801 North Senate Boulevard, Suite 230, Indianapolis, IN 46202.

## Abstract

**INTRODUCTION:** Hypoxemia in Severe Acute Respiratory Syndrome due to Novel Coronavirus of 2019 (SARS-CoV-2) is mediated by severe inflammation that may be mitigated by corticosteroids. We evaluated pattern and effects of corticosteroid use in these patients during an early surge of the pandemic.

**METHODS:** Observational study of 136 SARS-CoV-2 patients admitted to the Intensive care Unit between March 1 and April 27, 2020 at a tertiary care hospital in Indianapolis, USA.

Statistical comparison between cohorts and dosing pattern analysis was done. Outcome measures included number of patients requiring intubation, duration of mechanical ventilation, length of ICU stay and inpatient mortality.

**RESULTS:** Of 136 patients, 72 (53%) received corticosteroids. Groups demographics: Age (60.5 vs. 65; *p* .083), sex (47% male vs. 39% female; *p* .338) and comorbidities were similar. Corticosteroid group had increased severity of illness: PaO2/FiO2 (113 vs. 130; *p* .014) and SOFA (8 vs. 5.5; *p* < .001). Overall mortality (21% vs. 30%; *p* .234) or proportion of patients intubated (78 vs. 64%; *p* .078) was similar. Mortality was similar among mechanically ventilated (27% vs. 15%; *p* .151) however there were no deaths among patients who were not mechanically ventilated and received corticosteroids (0% vs. 57%; *p* <.001). Early administration (within 48 hours) showed decrease in proportion of intubation (66% vs. 87 vs. 100%; *p*.045), ICU days (6 vs., 16 vs. 18; *p* <.001), and ventilator days (3 vs. 12 & 14; *p* <.001). 45% received methylprednisolone.

**CONCLUSION:** Corticosteroids were used more frequently in SARS CoV-2 patients with higher severity of illness. Early administration of corticosteroids improved survival in non-mechanically ventilated patients; decreased ICU stay and may have prevented intubation.

## INTRODUCTION

The pandemic of a novel coronavirus originating in 2019 (COVID-19) that causes SARS-CoV-2 has affected millions worldwide resulting in hundreds of thousands of deaths since December 2019 [1].

In severe COVID-19, hypoxemic respiratory failure is characterized by bilateral lung infiltrates causing an Acute Respiratory Distress Syndrome (ARDS) by Berlin criteria [2, 17]. This respiratory failure is the cause of death in 70% of fatal COVID-19 cases [3].

The pathogenesis of SARS-CoV-2 infection and resulting clinical picture is a result of invasion of the respiratory and alveolar epithelium by this RNA betacoronavirus followed by an exaggerated host inflammatory response characterized by elevated ferritin, C-reactive protein and interleukin-6 (IL-6) among other pro-inflammatory substances [4]. The dysregulated host response ranges from a self-limiting local immune reaction elicited by cytokines released by macrophages and monocytes [5] to a severely dysfunctional immune response, which can cause severe pulmonary and systemic manifestations. In its severe form, a surge of pro-inflammatory cytokine release occurs similar to observations made during the original SARS-CoV and Middle East Coronavirus Respiratory Syndrome (MERS) epidemics [6]. The ensuing clinical picture therefore ranges from mild symptoms to severe hypoxemia due to heterogeneous, extensive and progressive disease in lungs often accompanied by elevated markers of inflammation and multiple organ dysfunction [7].

In addition to anti-viral treatments, anti-inflammatory and immune-modulating therapies are therefore being employed to ablate the cytokine storm of severe COVID-19. Pharmacological therapies targeting different steps of this inflammatory cascade are being investigated for use in COVID-19 patients. Therapies such as IL-6 inhibitors or other monoclonal antibodies, immunoglobulins and convalescent plasma aim to counter this pro-inflammatory milieu [8]. However despite the availability of novel treatments, corticosteroids continue to be an attractive option due to their potent anti-inflammatory and anti-fibrotic properties as well as their known use in severe respiratory inflammatory conditions including severe ARDS [9]. Recent data from a randomized control trial and a meta-analysis indicate favorable outcomes of corticosteroids in severe ARDS [10. 11].

Several published guidelines since the declaration of the COVID-19 pandemic, suggested that corticosteroids not be used routinely to treat this infection but to exercise the option to use corticosteroids in severe ARDS based on clinical discretion [12—14]. However the same guidelines suggest using corticosteroids if the criterion for severe ARDS is met.

Consequently there is heterogeneity in patterns of corticosteroid prescription in ARDS due to viral pneumonias in general as well as specifically for SARS CoV-2. A recent meta-analysis of corticosteroid use in SARS, MERS and SARS CoV-2 suggests that half of the patients received these agents. The type, dosage and timing of initiation of corticosteroids are also variable among providers [15]. The preliminary report of the Randomized Evaluation of COVid-19 thERapY (RECOVERY Trial) shows significantly improved outcome with dexamethasone and therefore revived this discussion [16].

The purpose of this study was to observe dosing patterns and clinical outcomes among patients with severe hypoxemic respiratory failure of SARS CoV-2 who received corticosteroids during an early surge of cases in our region.

## MATERIALS AND METHODS

### Study Population

The Indiana University (IU) Institutional Review Board (IRB) approved the conduct of this study and deemed it exempt (IRB study number 2005800938). Informed consent was waived, and de-identified data were analyzed. All patients that tested positive for SARS CoV-2 infection at the laboratory at Indiana University Health-Methodist Hospital between March 1 and April 27, 2020 were evaluated in this study. Methodist Hospital is an 802-bed academic tertiary-care center with 120 ICU beds. Patients less than 18-years old, imprisoned patients and pregnant patients were excluded.

### Study Definitions

ARDS was defined according to the Berlin criteria [17]. In patients with low pulmonary compliance standard ARDS-Network strategy was used [18]. Standardized treatment protocols were applied generally. Patients not receiving invasive mechanical ventilation received supplemental oxygen.

Coexisting medical conditions, medications, clinical examination findings, laboratory results, diagnostic imaging, treatment, and outcomes were reviewed from electronic medical records.

Sequential Organ Failure Assessment (SOFA) was calculated for all patients with using the worst value for each physiologic variable collected within 24 hours of admission to the intensive care unit. PaO2 to FiO2 (P/F) ratio was calculated using arterial blood gas measurements on ICU admission or value closest to time of intubation when available. Testing for COVID-19 was obtained using the Control and Prevention’s (CDC) assay [19]. Testing occurred at the Indiana State Department of Health Laboratory from March 1 to March 19 and thereafter at the IU Health Pathology Laboratory.

### Corticosteroids

Enteral and parenteral corticosteroid use was analyzed. Parenteral formulations included methylprednisolone, hydrocortisone and dexamethasone while prednisone was the enteral corticosteroid in our analysis. Dosage calculated and converted by available standard formulae to prednisone equivalent dose for purposes of consistent and accurate data analysis.

### Data Analysis

All continuous variables were summarized using median and interquartile range and were compared between study groups using Mann-Whitney U test. Categorical variables were summarized using count and percent and were compared between study groups using the Pearson Chi-square test or Fisher’s exact test, as appropriate. For comparisons across the 3 sub-groups defined by timing of steroids, the Kruskal Wallis test and Fisher’s exact test were used. A p value of p ≤ 0.05 was considered statistically significant. Statistical analysis was performed using Microsoft Excel (Version 16.38; Microsoft, 2020).

## RESULTS

### Demographic and Clinical Characteristics

626 patients tested positive for SARS CoV-2. 490 were outpatient or non-ICU inpatients. The remaining 136 were admitted to the ICU and were included in the analysis. 72 (53%) patients received corticosteroids compared to 64 (47%) who did not during that time period (Figure 1).

**Figure 1.**
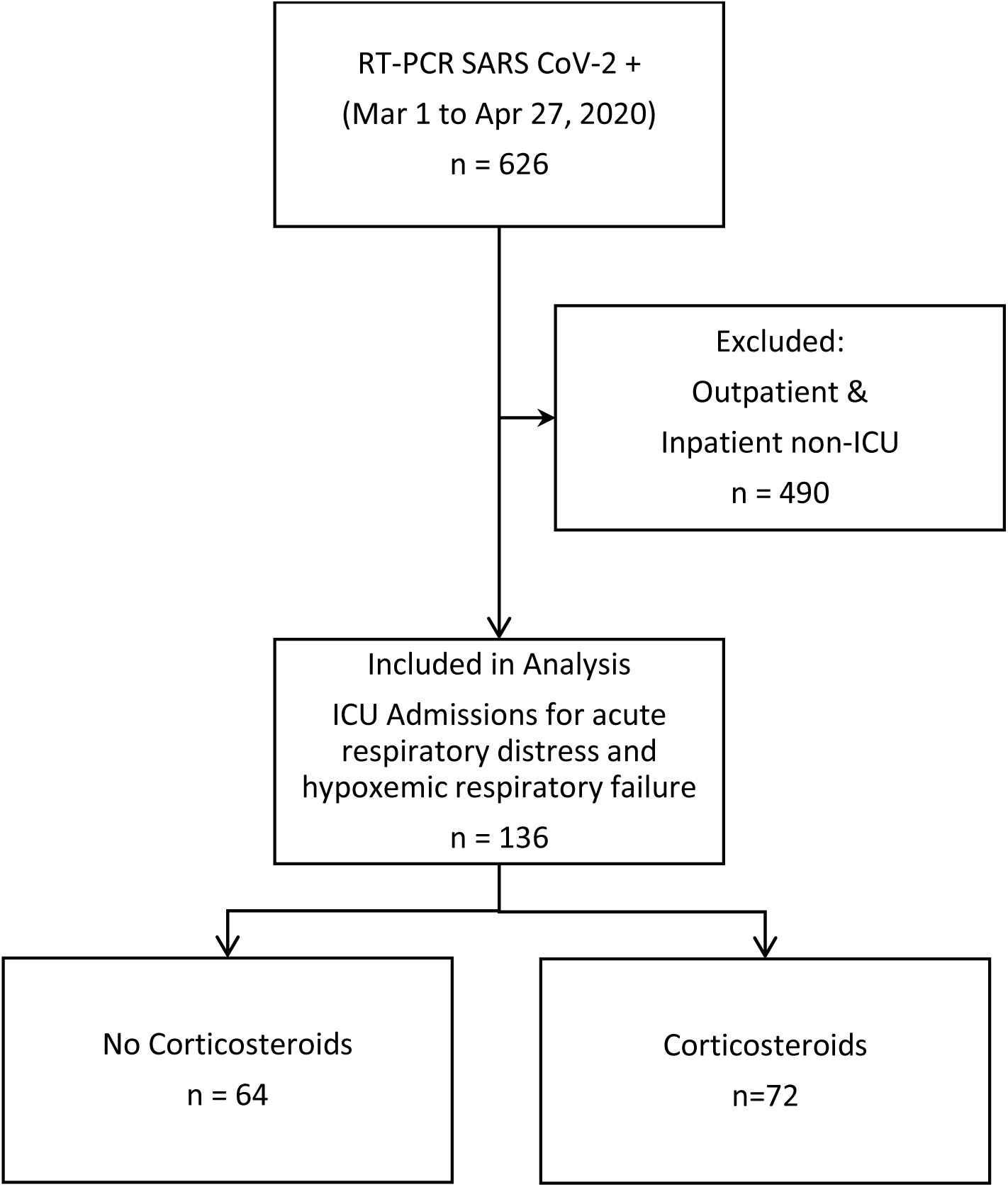
Study profile flow chart.

Demographic and clinical characteristics are shown in table 1. No significant difference is noted in age (60.5 vs. 65 years; p 0.083) or sex (47% male vs. 39% female; p 0.338) between the two groups. Comorbidities were similar including prevalence of COPD (8% vs. 14%; p 0.289) and asthma (14% vs. 9%; p 0.412). Presentation was similar: days of duration (6.5 vs. 7; p 0.218), respiratory symptoms (93% vs. 88%; p 0.271) and fevers/chills (67% vs. 59%; p 0.379). D-dimer values were higher in the non-steroid group (440 vs.686 ng/mL; p 0.003), LDH was higher in the steroid group (466 vs. 421 U/L; p 0.011). Ferritin and CRP were similar. The corticosteroid group was more severely ill: vasopressor use (65% vs. 41%; p 0.003), need for extracorporeal membrane oxygenation (7% vs. 0%; p 0.032), P/F ratio (113 vs. 130; p 0.014), SOFA score (8 vs. 5.5; p < 0.001) and ICU days (13.5 vs. 7; p <0.001) were worse in that cohort.

**Table 1:**
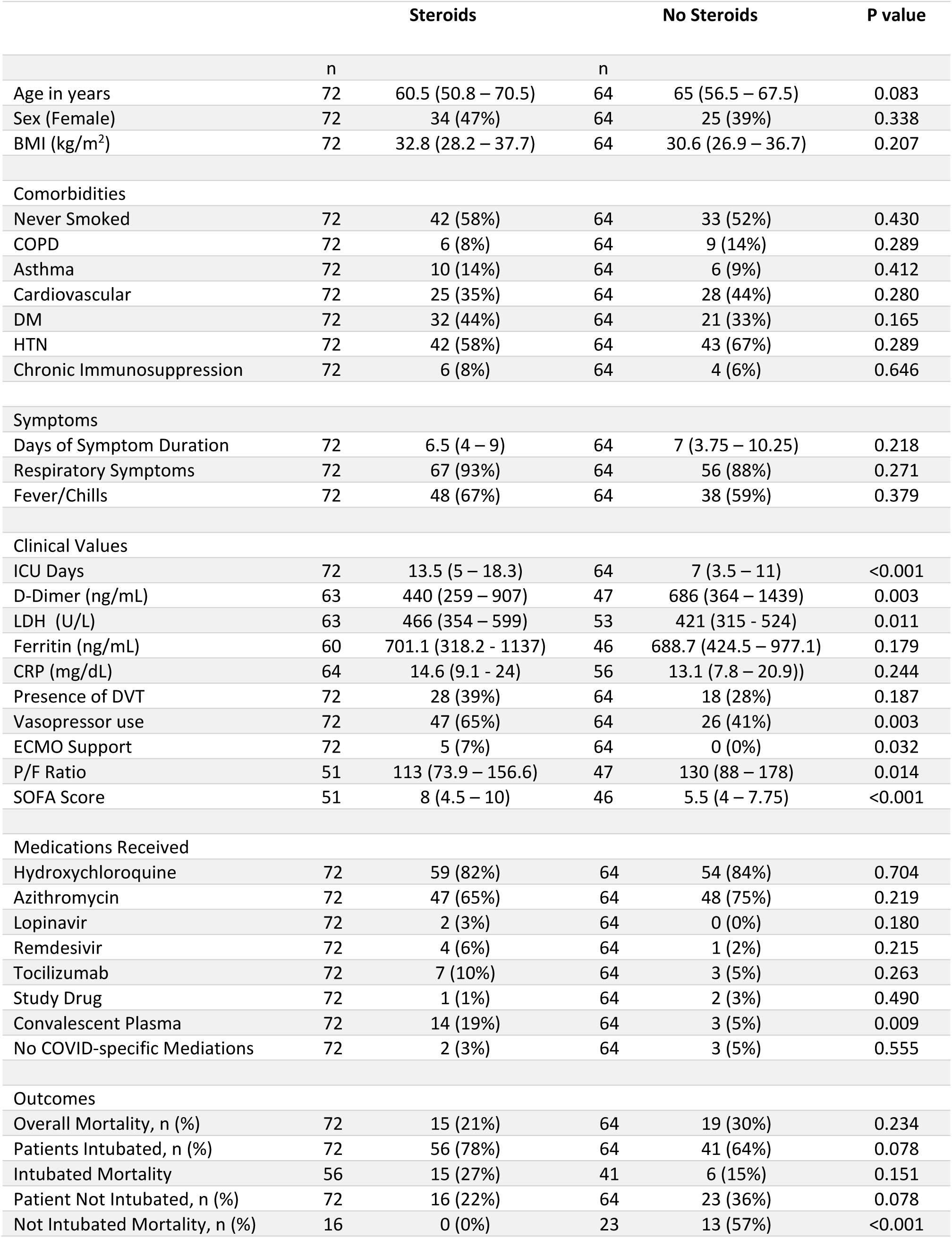
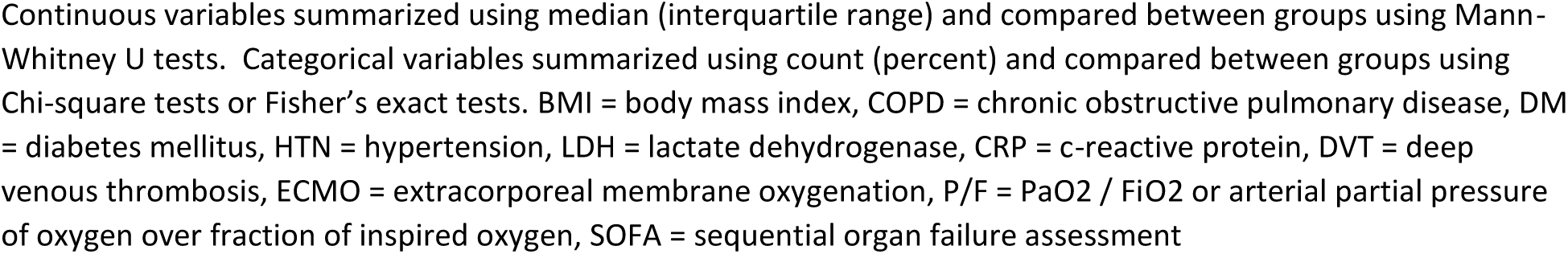
Demographics, comorbidities, prior symptoms, clinical values, medications received and outcome data.

|  | Steroids |  | No Steroids |  | P value |
| --- | --- | --- | --- | --- | --- |
|  | n |  | n |  |  |
| Age in years | 72 | 60.5 (50.8 – 70.5) | 64 | 65 (56.5 – 67.5) | 0.083 |
| Sex (Female) | 72 | 34 (47%) | 64 | 25 (39%) | 0.338 |
| BMI (kg/m<sup>2</sup>) | 72 | 32.8 (28.2 – 37.7) | 64 | 30.6 (26.9 – 36.7) | 0.207 |
| Comorbidities |  |  |  |  |  |
| Never Smoked | 72 | 42 (58%) | 64 | 33 (52%) | 0.430 |
| COPD | 72 | 6 (8%) | 64 | 9 (14%) | 0.289 |
| Asthma | 72 | 10 (14%) | 64 | 6 (9%) | 0.412 |
| Cardiovascular | 72 | 25 (35%) | 64 | 28 (44%) | 0.280 |
| DM | 72 | 32 (44%) | 64 | 21 (33%) | 0.165 |
| HTN | 72 | 42 (58%) | 64 | 43 (67%) | 0.289 |
| Chronic Immunosuppression | 72 | 6 (8%) | 64 | 4 (6%) | 0.646 |
| Symptoms |  |  |  |  |  |
| Days of Symptom Duration | 72 | 6.5 (4 – 9) | 64 | 7 (3.75 – 10.25) | 0.218 |
| Respiratory Symptoms | 72 | 67 (93%) | 64 | 56 (88%) | 0.271 |
| Fever/Chills | 72 | 48 (67%) | 64 | 38 (59%) | 0.379 |
| Clinical Values |  |  |  |  |  |
| ICU Days | 72 | 13.5 (5 – 18.3) | 64 | 7 (3.5 – 11) | <0.001 |
| D-Dimer (ng/mL) | 63 | 440 (259 – 907) | 47 | 686 (364 – 1439) | 0.003 |
| LDH (U/L) | 63 | 466 (354 – 599) | 53 | 421 (315 – 524) | 0.011 |
| Ferritin (ng/mL) | 60 | 701.1 (318.2 – 1137) | 46 | 688.7 (424.5 – 977.1) | 0.179 |
| CRP (mg/dL) | 64 | 14.6 (9.1 – 24) | 56 | 13.1 (7.8 – 20.9)) | 0.244 |
| Presence of DVT | 72 | 28 (39%) | 64 | 18 (28%) | 0.187 |
| Vasopressor use | 72 | 47 (65%) | 64 | 26 (41%) | 0.003 |
| ECMO Support | 72 | 5 (7%) | 64 | 0 (0%) | 0.032 |
| P/F Ratio | 51 | 113 (73.9 – 156.6) | 47 | 130 (88 – 178) | 0.014 |
| SOFA Score | 51 | 8 (4.5 – 10) | 46 | 5.5 (4 – 7.75) | <0.001 |
| Medications Received |  |  |  |  |  |
| Hydroxychloroquine | 72 | 59 (82%) | 64 | 54 (84%) | 0.704 |
| Azithromycin | 72 | 47 (65%) | 64 | 48 (75%) | 0.219 |
| Lopinavir | 72 | 2 (3%) | 64 | 0 (0%) | 0.180 |
| Remdesivir | 72 | 4 (6%) | 64 | 1 (2%) | 0.215 |
| Tocilizumab | 72 | 7 (10%) | 64 | 3 (5%) | 0.263 |
| Study Drug | 72 | 1 (1%) | 64 | 2 (3%) | 0.490 |
| Convalescent Plasma | 72 | 14 (19%) | 64 | 3 (5%) | 0.009 |
| No COVID-specific Mediations | 72 | 2 (3%) | 64 | 3 (5%) | 0.555 |
| Outcomes |  |  |  |  |  |
| Overall Mortality, n (%) | 72 | 15 (21%) | 64 | 19 (30%) | 0.234 |
| Patients Intubated, n (%) | 72 | 56 (78%) | 64 | 41 (64%) | 0.078 |
| Intubated Mortality | 56 | 15 (27%) | 41 | 6 (15%) | 0.151 |
| Patient Not Intubated, n (%) | 72 | 16 (22%) | 64 | 23 (36%) | 0.078 |
| Not Intubated Mortality, n (%) | 16 | 0 (0%) | 23 | 13 (57%) | <0.001 |

Continuous variables summarized using median (interquartile range) and compared between groups using Mann-Whitney U tests. Categorical variables summarized using count (percent) and compared between groups using Chi-square tests or Fisher's exact tests. BMI = body mass index, COPD = chronic obstructive pulmonary disease, DM = diabetes mellitus, HTN = hypertension, LDH = lactate dehydrogenase, CRP = c-reactive protein, DVT = deep venous thrombosis, ECMO = extracorporeal membrane oxygenation, P/F = $\text{PaO}_2 / \text{FiO}_2$ or arterial partial pressure of oxygen over fraction of inspired oxygen, SOFA = sequential organ failure assessment

COVID-specific therapy is also noted in Table 1. The majority of patients received concomitant hydoxychloroquine (83%) and azithromycin (70%).There was no significant difference except more convalescent plasma administration in the steroid-cohort (19% vs. 5%, p 0.009).

### Clinical Outcomes

There was no difference in overall mortality between the steroid and non-steroid receiving groups (21% vs. 30%; p 0.234). The proportion of intubated patients on invasive mechanical ventilation (78% vs. 64%; p 0.078) and mortality in intubated patients was similar (27% vs. 15%; p 0.015). However, the steroid-group had significantly decreased mortality among non-intubated patients (0% vs. 57%; p < 0.001). Outcomes are displayed graphically in Figure 2.

**Figure 2.**
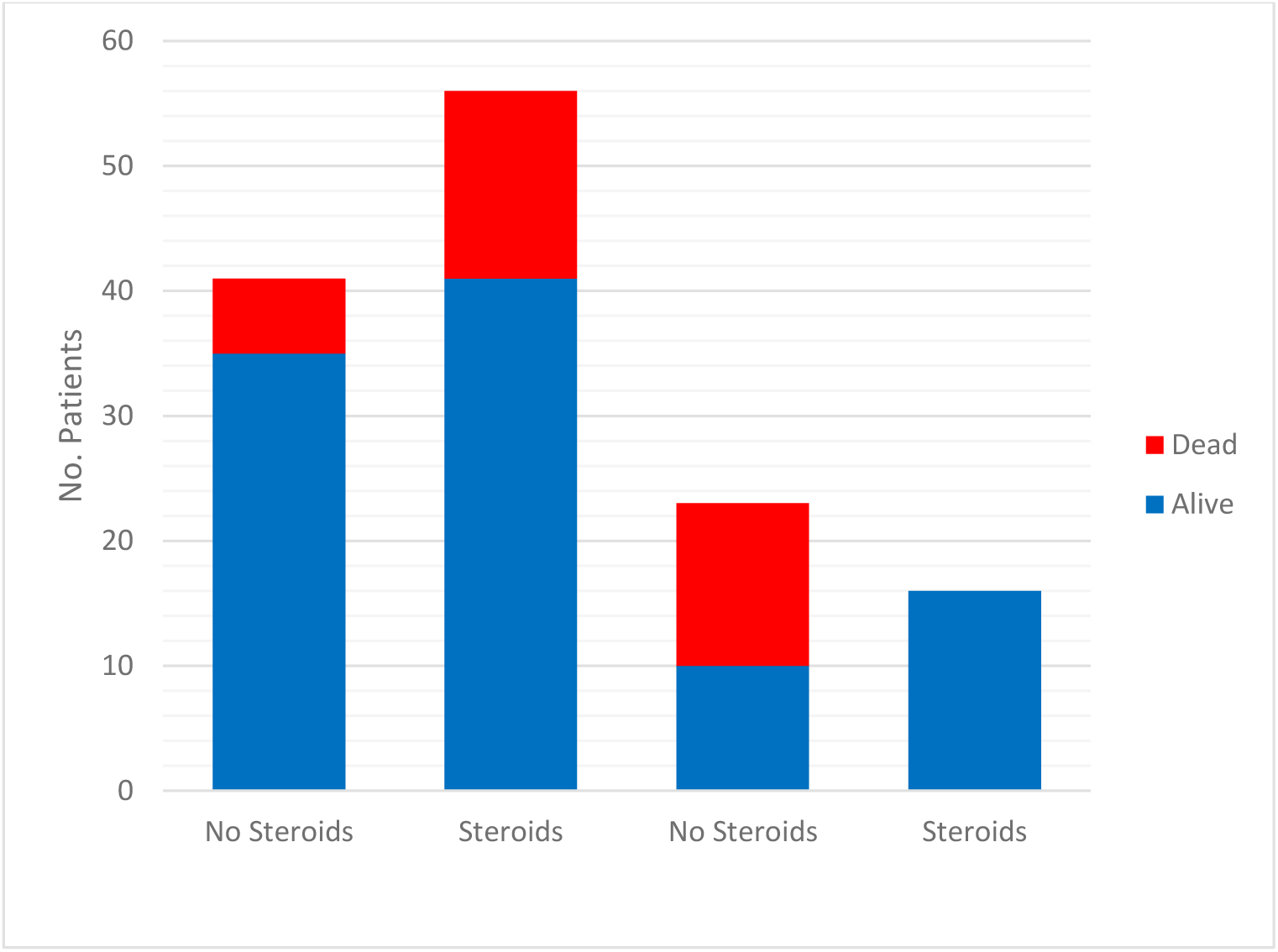
Graphical representation of respective deaths in patients receiving no steroids and those receiving steroids separated by whether they were on invasive mechanical ventilation (intubated) or oxygen supplementation without mechanical ventilation (non-intubated)

### Timing of Corticosteroid Administration

The steroid-group was further subdivided by timing of initial steroid dosing: *Early* if steroids were given less than 48 hours, *Mid* between 48 hours and 7 days, and *Late* greater than 7 days after ICU admission. Table 2 shows 35 patients in the *early*, 30 patients in the *mid*, and 7 patients in the *late* category. P/F ratio and SOFA score across these groups were similar (p 0.863 and p 0.831, respectively). Daily steroid dosing in prednisone equivalents and total steroid days was also not significantly different (p 0.458 and p 0.467, respectively). Significant difference was seen in proportion of intubation (66 vs. 87 vs. 100%; p 0.045), ICU days (6 vs. 16 vs. 18; p <0.001), and ventilator days (3 vs. 12 vs. 14; p <0.001). However, there was no difference in deaths between the three steroid timings (10 vs. 5 vs. 0; p 0.222).

**Table 2:** Summary of patients who receive steroids, broken down by when they received their first dose of steroids with respect to their ICU admission. *Early* patients received steroids within 48 hours of ICU admission, *Mid* patients received steroids between 48 hours and 7 days from admission. *Late* patients received their first dose of steroids greater than 7 days after ICU admission.

|  | Early (< 48hrs) | Mid (48hrs - 7 days) | Late (>7 days) | p-value |
| --- | --- | --- | --- | --- |
| N | 35 | 30 | 7 |  |
| P/F Ratio | 127 (72 – 157) | 104 (78 – 140) | 108 (76 – 164) | 0.863 |
| SOFA Score | 8 (4 – 10) | 8 (6 – 10) | 7 (4 – 10) | 0.831 |
| Daily Steroid Dosing in Prednisone Equivalents (mg) | 55 (32.8 – 100) | 56.7 (38.125 – 95.8) | 75 (51.68 – 100) | 0.458 |
| Days of Steroids | 7 (3.5 – 14.5) | 7.5 (6.25 – 17) | 9 (5 – 11) | 0.467 |
| Patients Intubated | 23 (66%) | 26 (87%) | 7 (100%) | 0.045 |
| Ventilator Days | 3 (0 – 11) | 12 (8 – 19) | 14 (13 – 17) | <0.001 |
| ICU Days | 6 (3 – 14.5) | 16 (11.5 – 22.75) | 18 (17.5 – 27) | <0.001 |
| Died, n (%) | 10 (29%) | 5 (17%) | 0 (0%) | 0.222 |

Continuous variables summarized using median (interquartile range) and compared between groups using Kruskal Wallis tests. Categorical variables summarized using count (percent) and compared between groups using Fisher's exact tests. IQR = interquartile range, ICU = intensive care unit

### Type and Dose of Corticosteroids

Distribution of corticosteroids given by total doses administered is noted in Figure 3. 717 doses of steroids were given: majority being intravenous methylprednisolone (325 doses, 45%). Most patients (90%) received steroids within the first 7 days, and the median daily dose administered was the equivalent of approximately 56 mg of prednisone. Prednisone, hydrocortisone, and dexamethasone were used at rates of 27%, 20%, and 8% respectively.

**Figure 3.**
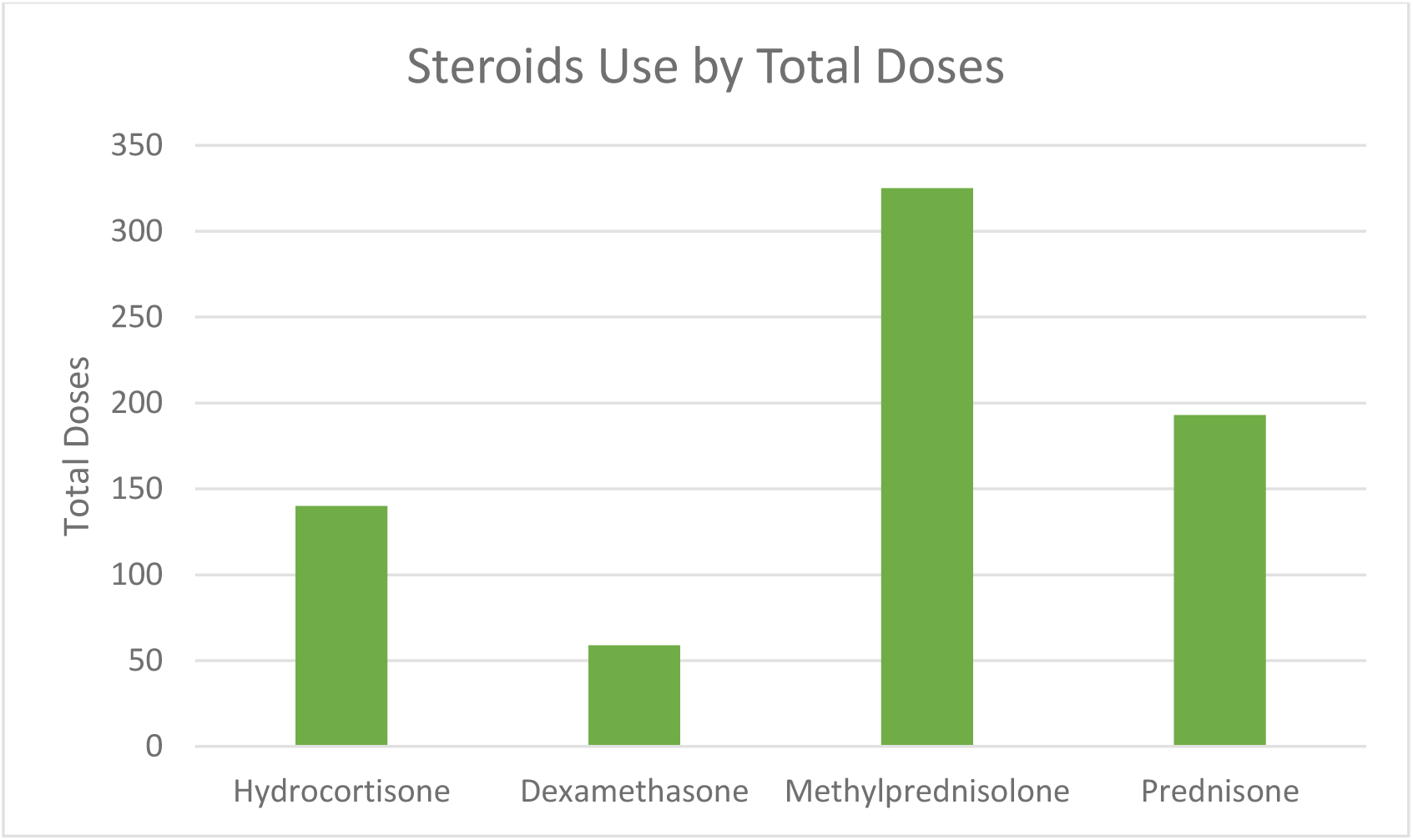
Distribution of corticosteroid type by total doses administered.

## DISCUSSION

We present our single center experience of dosing patterns and outcomes of corticosteroid use in severe hypoxemia associated with COVID-19 during an initial phase of the pandemic.

Although frequently debated, corticosteroids have been a treatment consideration of ARDS for decades [9, 20]. Mixed results due to small sample sizes, selection bias, patient heterogeneity, and time of initiation of treatment or duration of therapy have raised questions [20]. The ARDS Network’s large randomized trial of methylprednisolone versus placebo in 180 patients with ARDS of at least 7 days duration showed that while there was no survival benefit in the corticosteroid group, methylprednisolone increased the number of ventilator-free days and ICU-free days during the first 30 days [21]. A meta-analysis of data showed that prolonged administration of systemic steroids is associated with favorable outcomes and survival benefit when given before day 14 of ARDS [22]. Finally, in the 277 patient DEXA-ARDS trial by Villar J, et al, improvement in ventilator-free days and mortality in the dexamethasone administration arm of that trial (21% vs. 36%, *p*=0.004) was clearly demonstrated [11].

In addition to primary ARDS, there is data to suggest use of corticosteroids (such as hydrocortisone) in severe critical illnesses such as septic shock and severe pneumonia which are invariable linked to ARDS [23]. Positive data has been reported in ARDS related to septic shock especially when pneumonia is the underlying etiology [24].

Corticosteroid use in viral pneumonias such as influenza has been a source of controversy [25—28]. The guidelines for the administration of corticosteroids in SARS CoV-2 were mostly derived from the experience in previous epidemics of homologous coronaviruses namely SARS-CoV and MERS. Effect of corticosteroids in severe influenza is also included in these treatment guidelines from various societies that stated that corticosteroids should not be routinely used in SARS CoV-2 due to risk of delayed viral clearance, lack of mortality benefit and risk of longer hospitalizations due to side effects of these agents [13, 29—32].

However, corticosteroid use remains common in severe refractory hypoxemia to alleviate intense inflammation at the alveolar-capillary interface [33]. A meta-analysis of 11 studies showed that their use in SARS and MERS epidemics was more common in those with greater severity of illness [15]. Our data also confirms that corticosteroids were used in significantly greater frequency in those with more severe illness. This is expected as clinicians will tend to deploy a readily available therapy to target down-regulation of systemic and pulmonary inflammation.

A pooled data analysis of observational data from 4 studies and the preliminary report of the RECOVERY trial also favor corticosteroid use in COVID-19 [34, 16].

In RECOVERY, 2104/6425 patients were randomized to dexamethasone 6 mg daily for 10 days. Overall mortality benefit (21.6% vs. 24.6%; p < 0.001), mortality reduction of 35% (29.0% vs. 40.7%; p 0.003) in mechanically ventilated and 20% (21.5% vs. 25.0%; p 0.002) amongst non-mechanically ventilated is being reported in the preliminary findings [16]. Duration of illness of > 7 days before receiving dexamethasone seemed to have the greatest benefit.

Our data demonstrates heterogeneity in corticosteroid use, choice, dose, timing of initiation and duration of therapy in the early stage of this pandemic. We evaluated these trends and identified outcome variables based especially on the timing of initiation of corticosteroids after admission to the ICU. Our analysis presents a favorable outcome response especially if administered within 48 hours of ICU admission for prevention of intubation, duration of ICU-stay and mortality. This is consistent with data from the previous studies evaluating early steroid administration in ARDS. Notably, the mean time from intubation to randomization in the DEXA-ARDS study was two days [11]. Likewise, in the 2007 Meduri trial of early methylprednisolone for the treatment of ARDS, patients were enrolled within 72 hours of diagnosis. In this study of 92 patients, steroids were associated with a reduction in ventilator days, ICU days and mortality [35].

Most notably in our analysis, a significant mortality benefit was demonstrated in the corticosteroid-receiving patients who were not on invasive mechanical ventilation but receiving supplemental oxygen alone, as none of those in our cohort died. This may be similar to the preliminary report of the 85 patient GLUCOVID trial where survival benefit is being reported in non-mechanically ventilated patients that received a six-day course of methylprednisolone [36]. 45 % of our corticosteroid receiving patients in our study, received methylprednisolone and the duration ranged from seven to nine days.

Our data, however, does not demonstrate the same mortality benefit in patients receiving invasive mechanical ventilation. This may be due to high severity of illness of the patients at our center who received steroids. Several indicators including the SOFA score, P/F ratio and vasopressor use suggest that the corticosteroid group was a sicker patient population. Interestingly, as SOFA score is associated with rough estimate of mortality [37] and based on our scores, we expected mortality in the range of 30% in the steroid cohort vs. 20% in the non-steroid group, our corticosteroid group maintained outcomes similar to the non-steroid group and below the calculated expected mortality.

This analysis is limited as it is done retrospectively. We have limited sample size in a single center where there was variation in the described intervention of corticosteroids.

Also limiting, is co-existence of Covid-19 related therapies that were variably available during the study period. We recognize, that as the pandemic has progressed, we like others caring for these patients, have tried to expand our armamentarium as potential treatment options became available, in clinical trial or otherwise. As such, our internal standard care, varied with time. However, except for convalescent plasma in 14 patients in the corticosteroid group vs. 3 patients in the non-corticosteroid group, exposure to other potential therapies were similar between the groups. We do not yet fully understand the clinical impact convalescent plasma has on outcomes, and therefore any potential influence to our results. Lastly, we did not maintain a standard corticosteroid treatment plan. However, despite variation, our treatment patterns were similar to previously described protocols used for non-SARS COV-2 ARDS.

In summary, our data shows that despite the guidelines existent at that time, corticosteroids were used in more than 50% of patients with hypoxemic respiratory failure. Their use suggests a potential benefit, including that of decreased mortality, particularly in those that were not on invasive mechanical ventilation. Further clarification and evaluation of dosing schedules, timing of initiation and duration of treatment is warranted.

## CONCLUSION

Despite existing guidelines that recommended to the contrary, more than half of ICU patients with SARS CoV-2 received corticosteroids during the early wave of the COVID-19 pandemic.

In non-mechanically ventilated patients with hypoxemic respiratory failure due to SARS CoV-2, using corticosteroids was associated with significantly lower mortality. The early use of high dose corticosteroids for may have reduced number of ICU days.

Additionally, with similar baseline characteristics, use of corticosteroids in higher severity of illness was associated with similar mortality when compared with population without steroids and lower severity of illness.

## Data Availability

The datasets used and/or analyzed during the current study are available from the corresponding author on reasonable request.

## Author’s contribution

Acquisition, analysis, interpretation of data: OR, RAT, RMK, EMM, MKC, CLT, RK Conception, drafting, and revision of work: OR, RK, MKC, RAT, RML, EMM, SAP Final approval of work: OR, RAT, MKC, RMK, EMM, CLT, SAP, RK

## REFERENCES

1. World Health Organization. Coronavirus Disease (COVID-2019) Situation Report 162. Geneva, Switzerland: World Health Organization; June 30th, 2020.

2. Huang C, Wang Y, Li X, et al. Clinical features of patients infected with 2019 novel coronavirus in Wuhan, China. Lancet. 2020;395:497–506

3. Yang X, Yu Y, Xu J, et al. Clinical course and outcomes of critically ill patients with SARS-CoV-2 pneumonia in Wuhan, China: a single-centered, retrospective, observational study. Lancet Respir Med. 2020;8(5):475–481.

4. Tay MZ, Poh CM, Rénia L, MacAry PA, Ng LFP. The trinity of COVID-19: immunity, inflammation and intervention. Nat Rev Immunol. 2020;20(6):363–374

5. Law HK, Cheung CY, Ng HY, et al. Chemokine up-regulation in SARS-coronavirus-infected, monocyte-derived human dendritic cells. Blood. 2005;106(7):2366–2374.

6. Liu J, Zheng X, Tong Q, et al. Overlapping and discrete aspects of the pathology and pathogenesis of the emerging human pathogenic coronaviruses SARS-CoV, MERS-CoV, and 2019-nCoV. J Med Virol. 2020;92(5):491–494.

7. Xu Z, Shi L, Wang Y, et al. Pathological findings of COVID-19 associated with acute respiratory distress syndrome [published correction appears in Lancet Respir Med. 2020 Feb 25;:]. Lancet Respir Med. 2020;8(4):420–422.

8. Ye Q, Wang B, Mao J. The pathogenesis and treatment of the Ćytokine Storm’ in COVID-19. J Infect. 2020;80(6):607–613.

9. Wu C, Chen X, Cai Y, et al. Risk Factors Associated With Acute Respiratory Distress Syndrome and Death in Patients With Coronavirus Disease 2019 Pneumonia in Wuhan, China. JAMA Intern Med. 2020.

10. Villar J, Ferrando C, Martínez D, et al. Dexamethasone treatment for the acute respiratory distress syndrome: a multicentre, randomised controlled trial. Lancet Respir Med. 2020;8(3):267–276.

11. Sun S, Liu D, Zhang H, Zhang X, Wan B. Effect of different doses and time-courses of corticosteroid treatment in patients with acute respiratory distress syndrome: A metaanalysis. Exp Ther Med. 2019;18(6):4637–4644.

12. Russell CD, Millar JE, Baillie JK. Clinical evidence does not support corticosteroid treatment for 2019-nCoV lung injury. Lancet. 2020;395(10223):473–475.

13. Ni YN, Chen G, Sun J, Liang BM, Liang ZA. The effect of corticosteroids on mortality of patients with influenza pneumonia: a systematic review and meta-analysis]. Crit Care. 2019;23(1):99.

14. Alhazzani W, Møller MH, Arabi YM, et al. Surviving Sepsis Campaign: Guidelines on the Management of Critically Ill Adults with Coronavirus Disease 2019 (COVID-19). Crit Care Med. 2020;48(6):e440–e469.

15. Li H, Chen C, Hu F, et al. Impact of corticosteroid therapy on outcomes of persons with SARS-CoV-2, SARS-CoV, or MERS-CoV infection: a systematic review and meta-analysis. Leukemia. 2020;34(6):1503–1511.

16. RECOVERY Collaborative Group, Horby P, Lim WS, Emberson J, et al. Dexamethasone in Hospitalized Patients with Covid-19 - Preliminary Report. N Engl J Med. 2020;10.1056/NEJMoa2021436. doi:10.1056/NEJMoa2021436

17. Ferguson ND, Fan E, Camporota L, et al. The Berlin definition of ARDS: an expanded rationale, justification, and supplementary material. Intensive Care Med. 2012;38(10):1573–1582

18. Fan E, Del Sorbo L, Goligher EC, et al. An Official American Thoracic Society/European Society of Intensive Care Medicine/Society of Critical Care Medicine Clinical Practice Guideline: Mechanical Ventilation in Adult Patients with Acute Respiratory Distress Syndrome. Am J Respir Crit Care Med. 2017;195(9):1253–1263.

19. Centers for Disease Control and Prevention. Coronavirus disease 2019 (COVID-19). https://www.cdc.gov/coronavirus/2019-ncov/lab/rt-pcr-detection-instructions.html. Published 2020. Accessed 4/3/2020

20. Ruan SY, Lin HH, Huang CT, Kuo PH, Wu HD, Yu CJ. Exploring the heterogeneity of effects of corticosteroids on acute respiratory distress syndrome: a systematic review and metaanalysis. Crit Care. 2014;18(2):R63.

21. Steinberg KP, Hudson LD, Goodman RB, et al. Efficacy and safety of corticosteroids for persistent acute respiratory distress syndrome. N Engl J Med. 2006; 354(16):1671–1684

22. Tang BM, Craig JC, Eslick GD, Seppelt I, McLean AS. Use of corticosteroids in acute lung injury and acute respiratory distress syndrome: a systematic review and meta-analysis. Crit Care Med. 2009;37(5):1594–1603

23. Pastores SM, Annane D, Rochwerg B; Corticosteroid Guideline Task Force of SCCM and ESICM. Guidelines for the diagnosis and management of critical illness-related corticosteroid insufficiency (CIRCI) in critically ill patients (Part II): Society of Critical Care Medicine (SCCM) and European Society of Intensive Care Medicine (ESICM) 2017. Intensive Care Med. 2018;44(4):474–477.

24. Meijvis SC, Hardeman H, Remmelts HH, et al. Dexamethasone and length of hospital stay in patients with community-acquired pneumonia: a randomised, double-blind, placebo-controlled trial. Lancet. 2011;377(9782):2023–2030.

25. Yam LY, Lau AC, Lai FY, et al. Corticosteroid treatment of severe acute respiratory syndrome in Hong Kong. J Infect. 2007;54(1):28–39.

26. Chen RC, Tang XP, Tan SY, et al. Treatment of severe acute respiratory syndrome with glucosteroids: the Guangzhou experience. Chest. 2006;129(6):1441–1452. Ferreira FL, Bota DP, Bross A, et al. Serial evaluation of the SOFA score to predict outcome in critically ill patients.?JAMA. 2001;286(14):1754–8.

27. Arabi YM, Mandourah Y, Al-Hameed F, et al. Corticosteroid Therapy for Critically Ill Patients with Middle East Respiratory Syndrome. Am J Respir Crit Care Med. 2018;197(6):757–767.

28. Rodrigo C, Leonardi-Bee J, Nguyen-Van-Tam J, Lim WS. Corticosteroids as adjunctive therapy in the treatment of influenza. Cochrane Database Syst Rev. 2016;3:CD010406.

29. The Centers for Diseases Control and Prevention, CDC, USA Covid-19 treatment guidelines. https://files.covid19treatmentguidelines.nih.gov/guidelines/covid19treatmentguidelines.pdf9

30. Alhazzani W., Moller M., Arabi Y.M., Loeb M., Gong M.N., Rhodes A. Surviving Sepsis Campaign: guidelines on the management of critically ill adults with Coronavirus Disease 2019 (COVID-19) Intensive Care Med. 2020;46:854–887.

31. Chakraborty C, Sharma AR, Sharma G, Bhattacharya M, Lee SS. SARS-CoV-2 causing pneumonia-associated respiratory disorder (COVID-19): diagnostic and proposed therapeutic options. Eur Rev Med Pharmacol Sci. 2020;24(7):4016–4026.

32. Phua J, Weng L, Ling L, et al. Intensive care management of coronavirus disease 2019 (COVID-19): challenges and recommendations. Lancet Respir Med. 2020;8(5):506–517.

33. Meduri GU, Annane D, Chrousos GP, Marik PE, Sinclair SE. Activation and regulation of systemic inflammation in ARDS: rationale for prolonged glucocorticoid therapy. Chest. 2009;136(6):1631–1643.

34. Singh AK, Majumdar S, Singh R, Misra A. Role of corticosteroid in the management of COVID-19: A systemic review and a Clinician’s perspective. Diabetes Metab Syndr. 2020;14(5):971–978.

35. Meduri GU, Golden E, Freire AX, et al. Methylprednisolone infusion in early severe ARDS: results of a randomized controlled trial. Chest. 2007;131(4):954–963.

36. Corral L, Bahamonde A, Arnaiz de las Revillas F, et al. GLUCOCOVID: a controlled trial of methylprednisolone in adults hospitalized with COVID-19 pneumonia. June 18, 2020 https://www.medrxiv.org/content/10.1101/2020.06.17.20133579v1. preprint.

37. Vincent JL, de Mendonça A, Cantraine F, et al. Use of the SOFA score to assess the incidence of organ dysfunction/failure in intensive care units: results of a multicenter, prospective study. Crit Care Med. 1998;26(11):1793–800.

